# Relationship between neurochemical concentrations and neurofunctional measures in late-onset GM2 gangliosidosis

**DOI:** 10.1101/2022.09.11.22279836

**Authors:** D Rangaprakash, Akila Weerasekera, Olivia E Rowe, Christopher D Stephen, Florian S Eichler, Robert L Barry, Eva-Maria Ratai

## Abstract

Magnetic resonance spectroscopy (MRS) and functional MRI (fMRI), related through common biophysical bases, provide complementary information about brain function. The link between MRS and fMRI measures is of interest, especially in the ultra-rare, metabolic disease late-onset GM2 gangliosidosis (LOGG). Imaging studies on LOGG have been few and far between, with cerebellar atrophy and neurochemical impairments being the most prominent findings. However, it remains unknown as to how these neurochemical aberrations relate to neurofunctional characteristics. The goal of this study (7 LOGG, 7 age/sex matched controls) was to assess the relationship between MRS concentrations and fMRI measures derived from the same MRS ROI (cerebellum, thalamus, precuneus) in LOGG. To quantify the communication between MRS regions and rest of the brain, we employed graph measures estimated from resting-state fMRI functional connectivity. We found that one such measure, local efficiency, which quantifies the aggregate relationship between a MRS region and rest of the brain, was significantly associated with N-acetylaspartate (NAA) in the cerebellum and thalamus (*p*<0.05, FDR corrected). Poorer neuronal health, neuronal loss (NAA), and neuroinflammation (myo-inositol) were related to poorer cerebellum-brain communication. Likewise, reduced thalamus-brain communication was also associated with poorer neuronal health and longer disease duration (*p*=0.002). These findings hint at a model of impaired neurochemical concentrations in these regions, leading to aberrant communication between them and rest of the brain, which may exacerbate disease progression. Future research must replicate these findings in larger cohorts, and further investigate such abnormalities in the cerebellum, thalamus and precuneus in this ultra-rare neurological disease.

## 1. Introduction

Multimodal imaging of neurological disorders has gained traction in the past decade because they can provide complementary information and deeper insights into disease neurobiology. Magnetic resonance spectroscopy (MRS) and functional magnetic resonance imaging (fMRI) are two such modalities that provide complementary neurochemical and neurofunctional information, respectively. It is advantageous to assess how neurochemical and neurofunctional measures might be related in a given brain region, in addition to their individual impairments in a disease, to help us better understand the underlying neurobiology. This relationship is possible between MRS and fMRI because they have certain biophysical bases in common: some neurochemicals quantified through MRS modulate local hemodynamics, neurovascular coupling and neural activity (at a slower timescale), which are the bases of the blood oxygenation level dependent (BOLD) fMRI signal [1].

In this work, we studied the relationship between MRS neurochemical concentrations and certain fMRI-derived neurofunctional measures in late-onset GM2 gangliosidosis (LOGG), which is an ultra-rare form of GM2-gangliosidosis [2] that encompasses two distinguishable but closely related neurodegenerative disease clusters [3] called late-onset Sandhoff disease (LOSD) and late-onset Tay-Sachs disease (LOTS). LOGG neurobiology differs from early and juvenile-onset of GM2 gangliosidosis [4] [5]. LOGG is characterized by cerebellar ataxia, limb weakness, psychiatric issues, and cognitive impairments [5]. Case studies on LOGG have revealed cerebral atrophy [6], mild midbrain/brainstem atrophy [7], thalamic hypodensities [6], and white matter lesions [8]. Population-level neuroimaging studies have been few and far between in this disease. One MRS study found lower N-acetylaspartate (NAA) in occipital and thalamic white-matter [9], while cerebellar atrophy has been the most consistent observation in the few imaging studies so far [10] [11] [12] [13] [14] [15]. Our recently published study on LOGG found lower NAA (impaired neuronal health), higher myo-inositol (neuroinflammation), and atrophy in the cerebellum [16]. Despite progress made in this (limited) literature, it remains unclear how impairments observed in these separate imaging modalities are interrelated. Specifically, there is a large gap in our understanding of how neurochemical factors and neurofunctional characteristics are interrelated in metabolic diseases such as LOGG. This interrelationship is of interest because neurochemical factors drive the brain’s functional characteristics. To the best of our knowledge, this is the first multimodal MRS/fMRI study in this uncommon patient cohort.

Given this background, several fMRI measures were thoughtfully chosen by us. BOLD fMRI quantifies local hemodynamic response secondary to latent local neuronal activity. Among the various time series analysis techniques typically employed to understand the rich fMRI time series data, functional connectivity (FC) measures the strength of co-activation between pairs of brain regions. Resting-state FC has been popular with over 11,000 papers published to date (mostly in the past decade), and with 2000+ papers published in 2021 alone (source: PubMed). FC is relatively simple to implement and interpret, and is found to be a biomarker of most neurological and psychiatric disorders [17]. Yet one shortcoming of FC is that it is a bivariate measure, that is, FC computation only involves time series from two brain regions and does not consider other connections in the brain or how all the connections, taken together, are organized. In this study, our aim was to quantify the aggregate communication between regions having MRS data (MRS volume of interest or VOI) and the rest of the brain, so that this aggregate functional measure can be directly correlated with neurochemical concentrations of the MRS VOI. FC does not provide this information directly, and cannot be directly correlated with MRS concentrations either because FC represents a connection and not a region.

To overcome these shortcomings, we employed complex network modeling using graph theoretic techniques. It quantifies the architecture of whole-brain connectome and assesses the topology of the whole-brain FC network, making inferences on network structure. Graph measures obtained from FC network topology are capable of quantifying the functional role of a given MRS VOI in relation to its connectivity with the rest of the brain, providing one value per region and enabling us to directly compare (and correlate) them with corresponding MRS concentrations. Rubinov and Sporns [18] demonstrated the utility of graph measures in functional brain imaging, and this approach has found increasing application in brain imaging with over 1500 papers published to date (mostly in the past decade). Among various graph theoretic techniques, we focus here on certain nodal graph measures that quantify functional segregation (dense connectedness within subnetworks), which is necessary for optimal specialized processing in the brain. They measure whether other brain regions (i.e., nodes) connected to the MRS VOI (node) are connected among themselves, forming subnetworks with interconnected nodes. We chose this class of graph measures because they qualified the functional importance of the MRS VOI being investigated within the whole brain connectome, and also provided a single value per MRS VOI that can be directly compared with MRS neurochemical concentrations.

Studies assessing MRS-fMRI relationships have been uncommon in any disorder. A few earlier studies have considered the relationship between regional MRS and fMRI measures in both healthy and diseased populations. For instance, in healthy adults, one study reported a positive correlation between gamma-aminobutyric acid (GABA) and default mode network (DMN) fMRI deactivation, and a negative correlation between glutamate and the same [19]. Another study found a positive correlation between glutamate in the medial prefrontal cortex and fractional amplitude of low frequency fluctuations (fALFF), another fMRI-derived measure, in depression [20]. However, MRS-fMRI relationships have not been observed consistently across studies. For example, other studies found no correlation between MRS and BOLD activation [21] [22], although task fMRI BOLD activation findings might be task-specific and not generalizable. Also, studies often do not have a primary focus on the MRS-fMRI relationship, but rather only conflicting observations stemming from secondary/tertiary or supplemental analyses. To the best of our knowledge, no studies have previously assessed the relationships between graph measures and metabolite concentrations. The present study therefore seeks to advance our mechanistic understanding of LOGG neurobiology by assessing the interrelationship between complementary neurochemical and neurofunctional mechanisms – insights that cannot be gained by investigating either of these modalities in isolation. Given these sparse and mixed observations and the paucity of prior functional neuroimaging studies in LOGG, we did not develop *a priori* regional hypotheses about MRS-fMRI relationship but rather conducted an exploratory analysis assessing the relationship between fMRI graph measures of functional segregation and six neurochemicals available to us (choline, creatine, myo-inositol [mI], glutamate [Glx], NAA and mI/NAA ratio; defined in Methods section) in three VOIs: cerebellum, thalamus and precuneus. Our recent LOGG study reported abnormalities in mI and NAA [16], which is why we specifically considered the mI/NAA ratio in this study.

## 2. Methods

### 2.1. Participants

All participants provided informed consent and the MGH Institutional Review Board (IRB) approved the study. Seven adults with genetically confirmed diagnosis of LOGG were recruited either through the National Tay-Sachs and Allied Diseases Association (NTSAD) or Massachusetts General Hospital’s (MGH) Leukodystrophy Clinic. They were examined by a neurologist (ataxia specialist, C.D.S.). The diagnosis was defined by mutation analysis of HEXA and HEXB genes, or the absence (or near-absence) of HEX enzymatic activity (in white blood cells or the serum). Five of them had a diagnosis of the LOTS variant of LOGG, while two had the LOSD variant. Although there are some differences in these variants [16], we could not decipher them in our cohort due to their sample sizes; therefore, this study presents findings as a single LOGG cohort. Given the ultra-rare nature of LOGG (prevalence of about 0.0003% [23]), our sample size was small as an absolute number but respectable given that recruitment was challenging. Seven age and sex matched healthy controls were recruited, who were deemed healthy through self-report (no history of psychiatric or neurological illness). The inclusion criteria were as follows: all subjects had to be able and willing to undergo MRI (i.e., no contraindications), be 18 years of age or older, and able to provide informed consent.

The following clinical measures were administered by a board-certified neurologist (C.D.S.) [3]: Epworth sleepiness scale [25], Pittsburgh Sleep Quality Index (PSQI) [26], Eating Assessment Tool (EAT10, measures dysphagia) [27], Cerebellar Cognitive Affective Syndrome scale (CCAS) [29] (assess cognitive dysfunction seen in cerebellar disease); and clinical ataxia rating scales – Brief Ataxia Rating Scale (BARS) [28], LOGG severity [24], Friedreich Ataxia Rating Scale (FARS) [30], and Scale for the Assessment and Rating of Ataxia (SARA) [31]. BARS, SARA, and FARS scores were adjusted for clinical weakness to provide better ataxia severity assessment [3].

### 2.2. MRI data acquisition

All MRI data were obtained in a Siemens 3T Trio MRI scanner (Siemens Healthcare, Erlangen, Germany) in a single scan session using a 32-channel head coil at the Athinoula A. Martinos Center for Biomedical Imaging. MRS data were obtained using a single-voxel water-suppressed point-resolved spectroscopy (PRESS) sequence (TR/TE = 1700/30 ms, 128 averages, bandwidth=1200 Hz, vector size=1024) from three VOIs, viz. cerebellum (*x*=0, centered on cerebellar vermis, 2 × 2 × 2 mm^3^), left thalamus (2 × 2 × 1.5 mm^3^) and parietal cortex (specifically the precuneus at *x*=0, 2 × 2 × 2 mm^3^). Water suppression was enhanced through T1 effects (WET). The precuneus was initially chosen as a control region, while the cerebellum and thalamus were chosen because of prior evidence of their involvement in Tay-Sachs disease pathology [10] [11] [4]. MRS VOIs were shimmed locally using first and second order gradient double acquisition shims (GRE-shim) and first order manual shim. Identical water unsuppressed acquisitions (4 averages) were also acquired in each region. The final orientation, dimensions and rotation of every voxel was adjusted to maximize gray matter content. The final MRS VOI prescription was screen-captured in three orthogonal planes at the scanner console (Figure 1 in Rowe et al. [16] shows MRS VOI placement and spectra) (Supplemental Information section S1 shows MRS VOI centroids in each participant).

**Figure 1.**
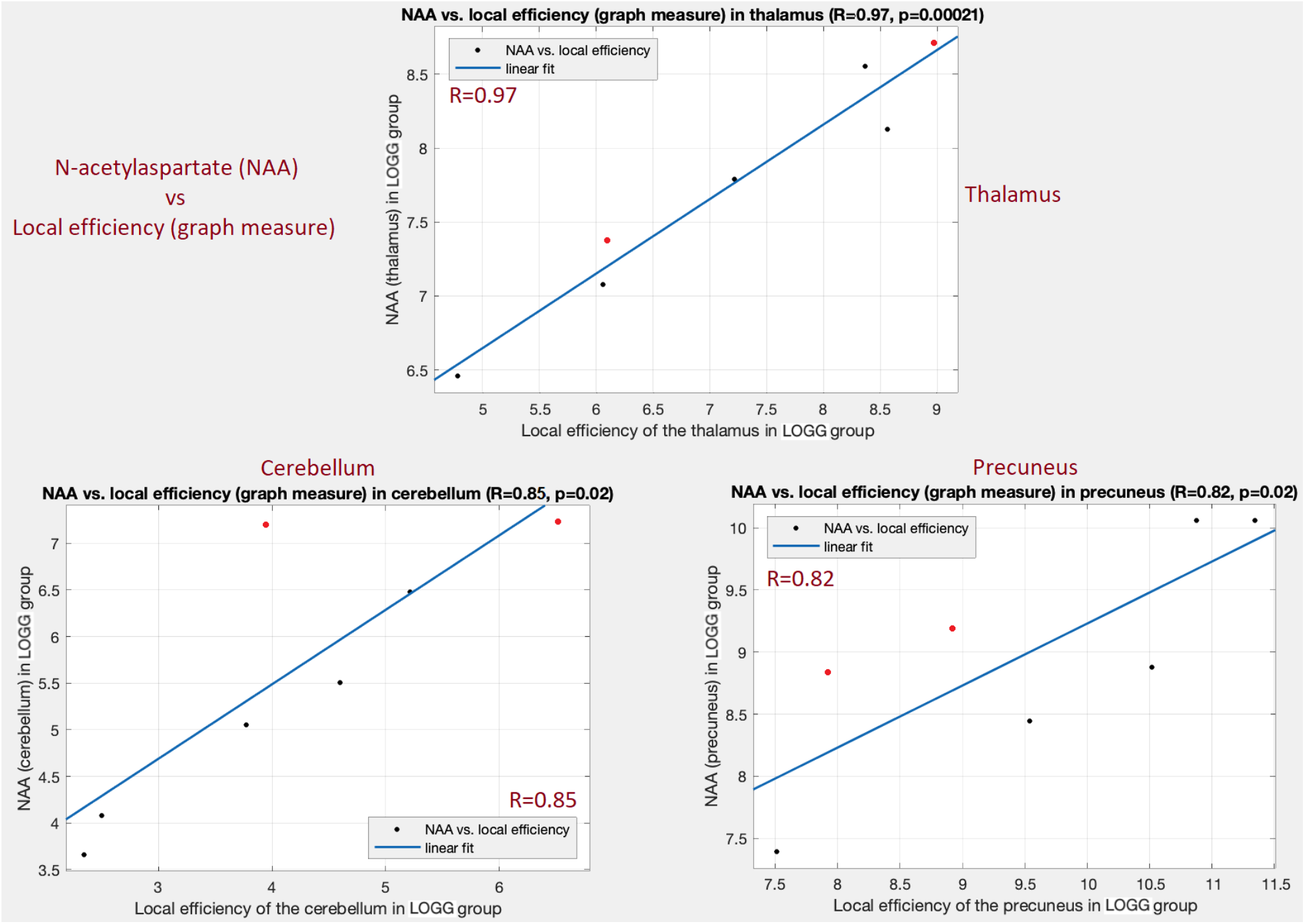
Significant associations between NAA and local efficiency graph measure (p<0.05, FDR corrected) in the LOGG group. Top: in thalamus (R = 0.97, p = 0.00021). Bottom-left: in cerebellum (R = 0.85, p = 0.0152). Bottom-right: in precuneus (R = 0.82, p = 0.0225). The two LOSD patients are visible as red points while the remaining blue points are LOTS patients.

FMRI data were obtained with the following scan parameters: 2D echo-planar imaging (EPI) sequence, TR/TE = 2000/30 ms, flip angle = 90°, field of view = 200×200 mm, voxel size = 2.5×2.5×4 mm^3^, 36 slices, scan duration = 10 min, whole brain coverage. Anatomical images, used to prescribe the MRS VOI, were obtained with the following parameters: 3D magnetization-prepared rapid acquisition of gradient echo (MPRAGE) sequence, TR/TE = 2530/1.64 ms, TI = 1200 ms, flip angle = 7°, field of view = 256×256 mm, voxel size = 1×1×1 mm^3^.

### 2.3. MRS data processing

MR spectra were fit with LCModel v6.3 [32] to obtain neurochemical levels in Institutional Units (IU), referenced to unsuppressed water from non-water-suppressed PRESS data. Absolute neurochemical levels were obtained per standard practice by dividing the integrated amplitude of the neurochemical signal by the integrated amplitude of water signal. Neurochemical values were discarded if LCModel’s percentage standard deviation exceeded 20% (Cramer-Rao lower bound). Data with obvious artifacts or poor quality were discarded (line width > 0.1 ppm, or signal-to-noise-ratio [SNR] < 5 calculated as the ratio of the NAA peak to the root-mean-squared value of the noise floor). The following neurochemicals were included: NAA, choline (Cho), creatine (Cr), myo-inositol (mI), and combined glutamate-glutamine (Glx). Each MRS VOI was transformed to the subject’s anatomical space and further segmented into white matter, gray matter, and cerebrospinal fluid (CSF) using Gannet toolkit v3.1 [33] and SPM12 [34]. Neurochemical estimates in each voxel were corrected for CSF variations using segmented tissue fractions [35]. The LCModel MRS spectra illustrated good signal-to-noise ratio (SNR = 22±8), shim and fit at <13% Cramer-Rao lower bound (≤ 20% is considered acceptable). These final MRS values from cerebellum, thalamus and precuneus regions were used in further analyses.

### 2.4. fMRI data pre-processing

FMRI data were pre-processed in MATLAB® using the CONN Toolbox [36], which is based on SPM12 [34]. The following pipeline was implemented: realignment, co-registration to MNI space, detection of outliers using Artifact detection Tools (ART) [37] (which also involved identifying high motion volumes with head motion above half of voxel size), and denoising. The denoising step involved regressing out the following nuisance variables: top ten principal components of CSF and white matter signals, twelve head motion parameters (3 rotation, 3 translation and their derivatives), and the outlier volumes identified with ART. Linear detrending and high-pass filtering (0.01 Hz) were thereafter performed. No subjects were excluded due to excessive head motion (aggregate motion > 1.25mm). Net head motion (mean frame-wise displacement [38]) was not statistically significant between groups (*p* = 0.29). Spatial smoothing was not conducted because a spatial averaging step was performed later during post-processing. Through visual inspection at every stage of analysis, we ensured accurate and reliable outputs to the best of our ability. All further analysis was performed in MATLAB® 2019b.

Given that fMRI is an indirect measure of neural activity, inter-subject and spatial variability of neurovascular coupling, measured by the hemodynamic response function (HRF) [39], has been found to confound fMRI connectivity [40] [41]. Additionally, connectivity impairments in mental disorders are confounded by HRF variability [42] [43]. Hence, deconvolution was performed on fMRI data [44] to estimate the HRF at each gray matter voxel and obtain HRF-variability-minimized 3D+time fMRI data, like in recent studies [45] [46] [47] [48] [49]. Thereafter, bandpass filtering was performed (0.01–0.1 Hz) on 3D+time deconvolved fMRI data using a 15^th^ order finite impulse response filter.

### 2.5. fMRI ROI selection procedure

Here we describe the procedure followed to define the fMRI regions of interest (ROIs). MRS data were obtained from three VOIs: cerebellum, thalamus and precuneus. We aimed to extract fMRI data from precisely the same MRS VOIs in each subject to directly compare MRS and fMRI data. MRS VOIs in each participant’s native anatomical space were transformed to the standard MNI space (nearest neighbor interpolation) using the transformation matrix obtained by co-registering the subject’s anatomical image to MNI space. The transformed MRS VOI centroids varied slightly across subjects (Supplemental Information Table S1). The median centroid locations in MNI space were: cerebellum (0, −2, −28), thalamus (−15, −16, 6), and precuneus (2, −44, 48). These were masked with the Harvard-Oxford 50% gray matter mask to obtain the final fMRI ROIs. Mean fMRI time series were extracted from each ROI for all subjects, giving us a time series with 300 points for each of cerebellum, thalamus and precuneus, which were then used in further analyses.

### 2.6. Associations between MRS concentrations and fMRI graph measures

The aim of graph analysis was to construct a whole-brain connectome involving each MRS VOI, measure the role of the MRS VOIs in the functional brain connectome and quantify their relationship with MRS concentrations. Whole-brain fMRI data were parcellated into 258 functionally homogenous ROIs, wherein 242 cortical ROIs were defined by the Power atlas [50] and 16 subcortical ROIs were defined by the Harvard-Oxford atlas [51]. Each MRS VOI (cerebellum, thalamus or precuneus) was added to the other brain ROIs to result in a total of 259 regions (this analysis was done separately for each of the three MRS VOIs). Any atlas ROI that overlapped spatially with the MRS VOI was excluded. Mean time series was obtained for each ROI by averaging time series from all gray matter voxels (50% mask) within the ROI. Then static FC was estimated between all pairs of 259 ROIs using Pearson’s product-moment correlation coefficient to generate a 259×259 connectivity matrix (separately for each of cerebellum, thalamus and precuneus) for each subject. Each FC value across subjects was statistically compared against null (i.e., FC=0) and all non-significant connections (*p*>0.05) were forced to 0. Absolute FC values were used hereafter.

Certain nodal graph measures were computed using the 259×259 connectome, giving us one value for each of the 259 ROIs. We only considered the value associated with the MRS VOI for further analysis. The following six graph measures were considered, which are briefly explained next (see [18] for a detailed rendering).

i. Node strength – average FC value of all connections associated with the MRS VOI (higher value implies overall higher connectivity between MRS VOI and the brain).
ii. Local efficiency – average reachability (inverse of shortest path length) between MRS VOI and other regions in the brain (higher value implies that other brain regions find it easier to communicate with the MRS VOI).
iii. Betweenness centrality – fraction of shortest paths in the entire network that go through the MRS VOI (higher value implies that the MRS VOI is important for communication between other pairs of brain regions, i.e., that it is an important mediator in the entire brain network).
iv. Eigenvector centrality – a measure of rich-club wherein the MRS VOI would have higher eigenvector centrality if it were well connected to other regions with higher eigenvector centrality (higher value implies that the MRS VOI is well connected to certain other important brain regions that drive network behavior).

Upon obtaining these nodal graph measures for the cerebellum, thalamus and precuneus VOIs across all subjects, we then assessed their associations with the six MRS concentrations within the LOGG group (*p*<0.05, corrected for multiple comparisons using the false discovery rate [FDR] method). The regressions were controlled for the ratio of gray matter to white matter volume within the MRS VOIs. In order to link *neurochemical-neurofunctional* relationships with LOGG impairments, group differences in these graph measures were assessed (*p*<0.05, FDR corrected). In the interest of future studies – considering the fact that LOGG is an ultra-rare disease leading to our modest sample size – we also report significant associations without multiple comparisons correction, and explicitly state the same where reported. Finally, we probed the association of significant graph measures with the clinical variables mentioned earlier (*p*<0.05, FDR corrected).

## 3. Results

Seven adults with genetically confirmed diagnosis of LOGG as well as 7 healthy age and sex matched controls were enrolled in this study. **Table 1** provides clinical measures and demographic information (comprehensively presented in an earlier study on the same cohort [3]). Age was not statistically significant between groups (*p* = 0.95, *Z* = −0.06, Wilcoxon rank-sum test). Group differences in MRS values and gray matter volume are reported in another study from our group on the same cohort [16]. Briefly, lower NAA and higher mI were observed only in the cerebellum, which were associated with certain clinical variables. No group differences in MRS values (but “trend level” observations in some cases) were noticed in thalamus and precuneus. Atrophy was observed in the cerebellum.

**Table 1.** Demographics and clinical variables. Apart from age and gender, all other measures pertain to the LOGG group alone.

| Variable | mean ± SD | Range |
| --- | --- | --- |
| <u>Disease demographics</u> |  |  |
| Age (LOGG) (years) | 42.9 ± 7.4 | 22 – 62 |
| Age (control) (years) | 42.7 ± 7.2 | 23 – 62 |
| Sex (control) | 4 M, 3 F | - |
| Sex (LOGG) | 4 M, 3 F | - |
| Disease duration (y) | 27.9 ± 4.5 | 12 – 39 |
| Age at disease onset | 15 ± 3.6 | 8 – 26 |
| <u>Clinical rating scales</u> |  |  |
| LOGG severity score | 7.1 ± 1.5 | 4 – 13 |
| SARA | 10.4 ± 1.9 | 4.5 – 16.5 |
| FARS | 33.1 ± 6.5 | 18 – 55.5 |
| BARS | 5.4 ± 2.3 | 1.5 – 15 |
| <u>Cognitive/psychiatric scales</u> |  |  |
| CCAS | 98.9 ± 5.9 | 76 – 114 |
| <u>Sleep scales</u> |  |  |
| ESS | 9.7 ± 3.1 | 2 – 21 |
| PSQI | 7.9 ± 3.2 | 2 – 20 |
| <u>Other scales</u> |  |  |
| EAT10 | 9 ± 4.3 | 0 – 22 |

BARS = brief ataxia rating scale; CCAS = cerebellar cognitive affective syndrome scale; FARS = Friedreich ataxia rating scale; EAT10 = eating assessment tool; SARA = Scale for the Assessment and Rating of Ataxia; PSQI = Pittsburgh sleep quality index; LOGG = late-onset GM2 gangliosidosis; ESS = Epworth sleepiness scale.

### 3.1 Graph analysis results

With group comparisons (LOGG vs. controls), local efficiency was significantly lower in LOGG compared to controls in the thalamus (*p* = 6.5×10^−4^, *T* = 4.57, Cohen’s *d* = 2.44), indicative of poorer thalamus-brain communication in LOGG. Without multiple comparisons correction, local efficiency was also significantly lower in LOGG in the precuneus (*p* = 0.034, *T* = 2.39, Cohen’s *d* = 1.28). Local efficiency was not different in the cerebellum (*p* = 0.35). Other graph measures did not differ between the groups in the three MRS VOIs (*p*>0.05).

Local efficiency was significantly positively associated with NAA in the thalamus in LOGG (R = 0.97, R^2^ = 0.94, *p* = 2.1×10^−4^) (**Table 2**). The regression fit (**Figure 1**) was found to be fully robust upon performing certain robustness tests: (i) there were no influential observations, that is, performing regression by leaving out one data point and repeating this for all possible combinations did not yield a single combination with non-significant association (i.e., all associations were significant even if a random data point were dropped), implying that outliers did not drive the association; (ii) the min and max *p*-values with leave-one-out regression were *p*_*min*_ = 0.0001 and *p*_*max*_ = 0.0039 (*p*<0.05); (iii) the residuals were homoscedastic (*p* = 0.27, Engle’s arch test for residual heteroscedasticity), ensuring that the homoscedasticity assumption of linear regression was not violated (i.e., data points were evenly scattered; homogeneity of variance).

**Table 2.** Significant associations between MRS concentrations and local efficiency (graph measure) in LOGG (p<0.05, FDR corrected). NAA and local efficiency were positively associated in all three regions, with the most prominent association in thalamus (R = 0.97, p = 0.00021). In the cerebellum, four associations were found, with a positive relationship with NAA and Glx and a negative one with mI and mI/NAA. Poorer cerebellum-brain communication was related to lower NAA and Glx and elevated mI. None of these associations were observed in controls. NAA = N-acetyl aspartate, mI = myo-inositol, Glx = glutamate + glutamine.

|  | Graph measure | MRS measure | R-value | R<sup>2</sup> value | p-value |
| --- | --- | --- | --- | --- | --- |
|  | Cerebellum |  |  |  |  |
| 1 | Local efficiency | NAA | 0.85 | 0.72 | 0.0152 |
| 2 |  | ml | −0.87 | 0.76 | 0.0107 |
| 3 |  | ml / NAA | −0.84 | 0.71 | 0.0164 |
| 4 |  | Glx | 0.88 | 0.77 | 0.0084 |
|  | Thalamus |  |  |  |  |
| 5 | Local efficiency | NAA | 0.97 | 0.94 | 2.1 × 10<sup>−4</sup> |
|  | Precuneus |  |  |  |  |
| 6 | Local efficiency | NAA | 0.82 | 0.67 | 0.0225 |

In LOGG, local efficiency was also significantly positively associated with NAA in the cerebellum (R = 0.85, R^2^ = 0.72, *p* = 0.02) and precuneus (R = 0.82, R^2^ = 0.67, *p* = 0.02). The cerebellum regression fit was moderately robust: there were 3 influential observations (*p*_*min*_ = 1.1×10^−4^, *p*_*max*_ = 0.07), residuals were homoscedastic (*p* = 0.45). The precuneus regression fit was moderately robust: 3 influential observations (*p*_*min*_ = 0.021, *p*_*max*_ = 0.15), residuals were homoscedastic (*p* = 0.52). **Figure 1** illustrates the three significant associations observed between NAA and local efficiency. In this and subsequent figures, LOSD patients are shown in a different color (red). It appears evident that these associations were not driven by differences between LOSD and LOTS, but we could not statistically demonstrate this because there were only 2 LOSD patients. None of these associations were significant in controls (*p*>0.50).

In the cerebellum, local efficiency was also associated with two other neurochemicals (**Table 2**) other than that with NAA presented earlier. It was associated with (**Figure 2**) myo-inositol (R = −0.87, R^2^ = 0.76, *p* = 0.0107), mI/NAA ratio (R = −0.84, R^2^ = 0.71, *p* = 0.0164), and Glx (R = 0.88, R^2^ = 0.77, *p* = 0.0084). The myo-inositol association was moderately robust: 1 influential observation (*p*_*min*_ = 0.0013, *p*_*max*_ = 0.06), homoscedastic residuals (*p* = 0.25). The mI/NAA association was fully robust: no influential observations (*p*_*min*_ = 8.9 × 10^−4^, *p*_*max*_ = 0.049), homoscedastic residuals (*p* = 0.68). The Glx association was moderately robust: 2 influential observations (*p*_*min*_ = 0.006, *p*_*max*_ = 0.11), homoscedastic residuals (*p* = 0.17). Overall, better cerebellum-brain communication was related to higher NAA and Glx, and to lower mI and mI/NAA. Since long-distance cerebellum-brain communication is central to the cerebellum’s role, it is understandable that local efficiency was related to neurochemical concentrations here. None of these associations were significant in controls, although associations between local efficiency and Glx were close to trend level in cerebellum (*p* = 0.09), thalamus (*p* = 0.13) and precuneus (*p* = 0.19).

**Figure 2.**
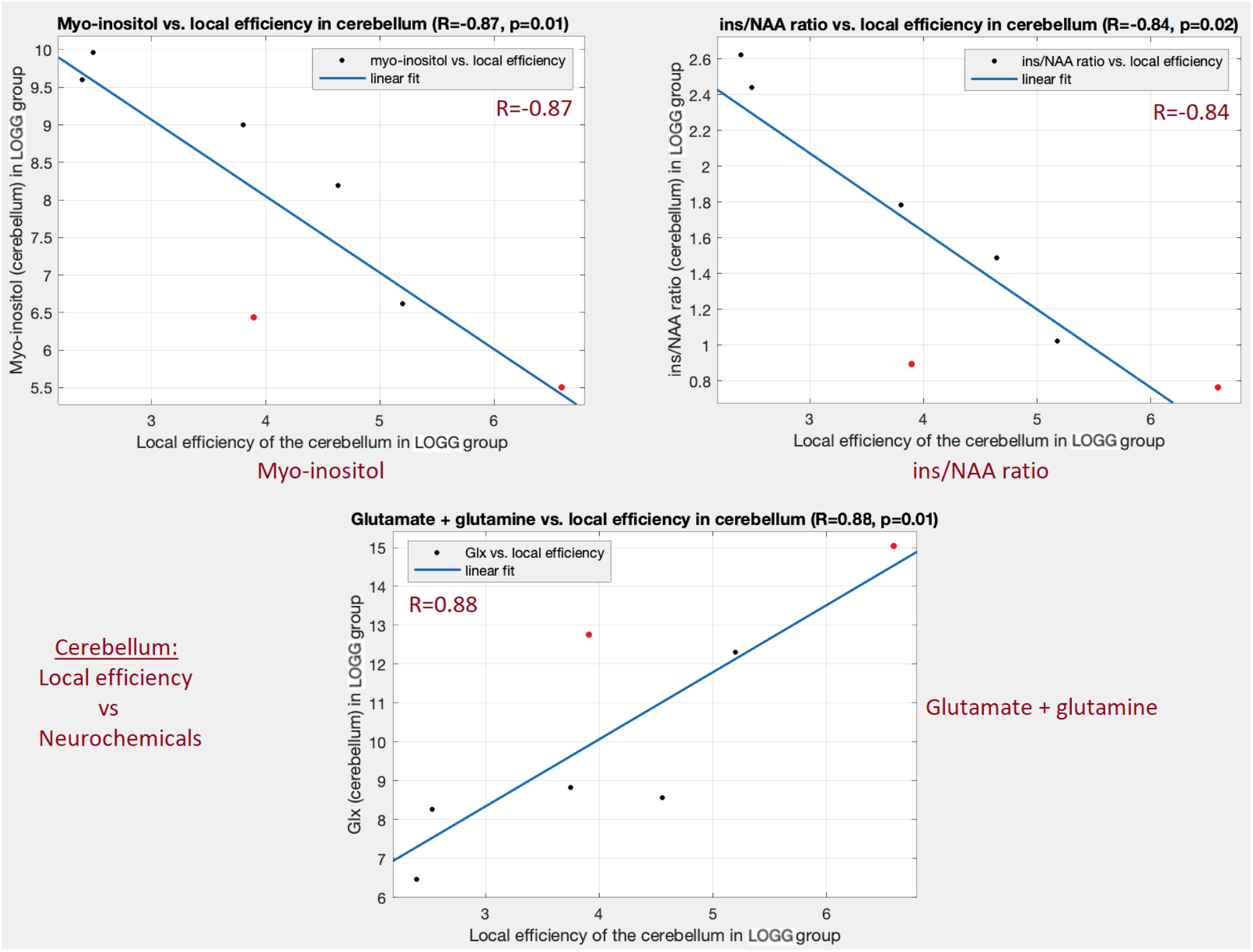
Significant associations between local efficiency and some neurochemicals in the cerebellum (p<0.05, FDR corrected) in the LOGG group. Top-left: with myo-inositol (mI) (R = −0.87, p = 0.0107). Top-right: with mI/NAA ratio (R = −0.84, p = 0.0164). Bottom: with Glx (glutamate + glutamine) (R = 0.88, p = 0.0084). Local efficiency in cerebellum was also associated with NAA as shown in Figure 1. The two LOSD patients are visible as red points.

Given that four neurochemicals were associated with local efficiency in the cerebellum in LOGG, we sought to understand the aggregate relationship of the four neurochemicals (taken together) with local efficiency. We employed partial least squares (PLS) regression [52] to this effect. We found that the aggregate of NAA, mI, mI/NAA and Glx explained 74% variance in local efficiency (R = 0.86, R^2^ = 0.75, *p* = 0.012), which was about the same as the variance explained by each metabolite individually (68%, 77%, 76% and 74% respectively). This hints towards a common substrate underlying these relationships.

Apart from the local efficiency measure, centrality measures (eigenvector and betweenness centralities) were associated with both NAA and creatine in the thalamus in LOGG (without multiple comparisons correction) (**Table 3**). Since the thalamus is a centrally located brain communication center [53], it is understandable that centrality measures were associated with neurochemical concentrations here. We employed PLS regression to assess the aggregate relationship between the two centrality measures and NAA/creatine (in both regions) and found that 74% variance in the aggregate of NAA and creatine was explained by the aggregate of the two centrality measures (R = 0.86, R^2^ = 0.73, *p* = 0.014), which was only about 8% higher than that of individual associations between these variables. NAA and creatine, taken together, might drive centrality characteristics of the thalamus in LOGG.

**Table 3.**
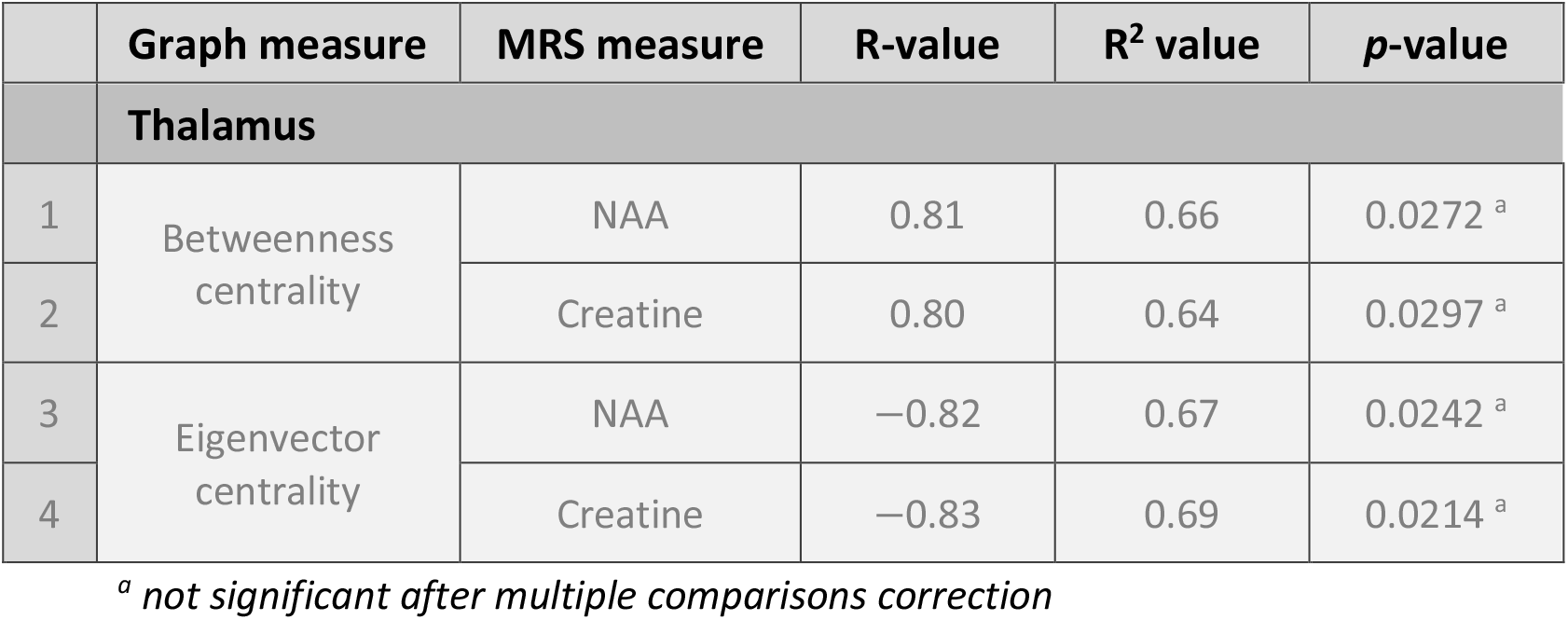
Significant associations between MRS concentrations and centrality graph measures in the thalamus, without multiple comparisons correction (none were significant after correction). Betweenness centrality and eigenvector centrality were each associated with both NAA and creatine. Betweenness centrality was positively related while eigenvector centrality was negatively related. NAA = N-acetyl aspartate, creatine = Cr+PCr (total creatine + phosphocreatine).

|  | Graph measure | MRS measure | R-value | R<sup>2</sup> value | p-value |
| --- | --- | --- | --- | --- | --- |
|  | **Thalamus** |  |  |  |  |
| 1 | Betweenness centrality | NAA | 0.81 | 0.66 | 0.0272 <sup>a</sup> |
| 2 |  | Creatine | 0.80 | 0.64 | 0.0297 <sup>a</sup> |
| 3 | Eigenvector centrality | NAA | −0.82 | 0.67 | 0.0242 <sup>a</sup> |
| 4 |  | Creatine | −0.83 | 0.69 | 0.0214 <sup>a</sup> |

<sup>a</sup> not significant after multiple comparisons correction

Node strength was not significantly associated with any neurochemical in any region in either group (see Supplemental Information section S2). In the interest of future studies, we additionally performed certain tertiary analyses with the strength of low-frequency BOLD fluctuations (fALFF) and neurovascular coupling (HRF), which are not part of our study design or conclusions; we direct the readers to Supplemental Information section S3. Age did not impact our results (see Supplemental Information section S4).

### 3.2. Associations between fMRI measures and clinical variables

Local efficiency in the thalamus was significantly negatively associated with disease duration (R = −0.94, R^2^ = 0.88, *p* = 1.7×10^−3^) (**Table 4**). Longer disease duration was related to overall poorer communication between the thalamus and rest of the brain, implying that having the disease for longer reduces thalamo-cortical connectivity. The regression fit (**Figure 3**) was fully robust: no influential observations (*p*_*min*_ = 0.0017, *p*_*max*_ = 0.0358), homoscedastic residuals (*p* = 0.95). Without multiple comparisons correction, local efficiency in the cerebellum was also associated with age of onset of LOGG (**Table 4**).

**Table 4.** Significant associations between graph measures and clinical variables. The association of local efficiency and centrality measures in the thalamus with disease duration were significant with multiple comparisons correction, while the association between age of onset and local efficiency in the cerebellum was not (in gray).

|  | Graph measure | Clinical measure | R-value | R<sup>2</sup> value | p-value |
| --- | --- | --- | --- | --- | --- |
| **Cerebellum** |  |  |  |  |  |
| 1 | Local efficiency | Age of onset | 0.80 | 0.64 | 0.0315 <sup>a</sup> |
| **Thalamus** |  |  |  |  |  |
| 2 | Local efficiency | Disease duration | -0.94 | 0.88 | 0.0017 |
| 3 | Betweenness centrality |  | -0.85 | 0.72 | 0.0151 |
| 4 | Eigenvector centrality |  | 0.89 | 0.79 | 0.0076 |

<sup>a</sup> not significant after multiple comparisons correction

**Figure 3.**
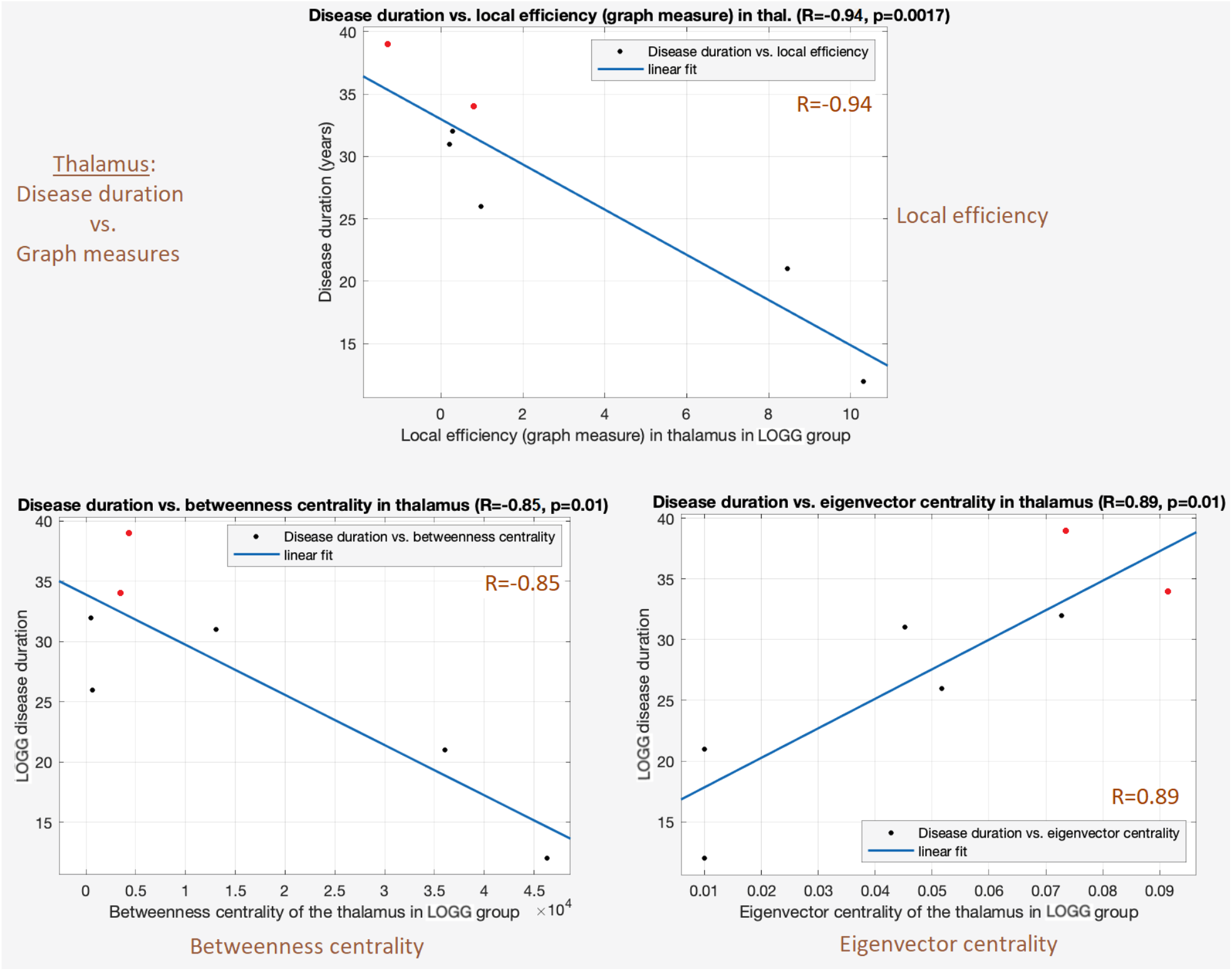
Significant associations between disease duration and graph measures in the thalamus (p<0.05, FDR corrected) in the LOGG group. Top: disease duration vs. local efficiency (R = −0.94, p = 0.0017). Bottom-left: disease duration vs. betweenness centrality (R = −0.85, p = 0.0151). Bottom-right: disease duration vs. eigenvector centrality (R = 0.89, p = 0.0076). Longer disease duration was related to poorer thalamo-cortical communication. The two LOSD patients are visible as red points.

Centrality measures in the thalamus were also significantly associated with LOGG disease duration (**Table 4**). The betweenness centrality regression fit was moderately robust: 1 influential observation (*p*_*min*_ = 0.0046, *p*_*max*_ = 0.16), homoscedastic residuals (*p* = 0.31). The eigenvector centrality regression fit was moderately robust: no influential observations (*p*_*min*_ = 0.011, *p*_*max*_ = 0.039), residuals were barely homoscedastic (*p* = 0.071). **Figure 3** illustrates the three significant associations observed between thalamic graph measures and disease duration.

## 4. Discussion

The goal of this study was to assess the relationship between MRS concentrations and fMRI graph measures derived from the same MRS VOIs in LOGG. Given the paucity of prior literature on this topic, our exploratory analysis probed associations between six neurochemicals and four graph measures (local efficiency, node strength, eigenvector centrality, betweenness centrality), as well as their group differences and their associations with clinical variables, with the hope of developing a better understanding of neurochemical and neurofunctional impairments in LOGG, and the relationship between the two.

We identified certain MRS-fMRI relationships in LOGG, specifically (i) efficiency of cerebellum-brain communication (local efficiency) was related to four neurochemicals in the cerebellum including NAA, (ii) local efficiency was related to NAA in all three regions, and (iii) centrality of thalamus (i.e., importance of the thalamus in cortical-cortical communication) was related to NAA and creatine (without multiple comparisons correction). Given that these observations were exclusive to the LOGG group and not noticed in the well-matched control group, they were likely driven by the underlying disease pathology. Taken together, our study found that certain neurochemical mechanisms (or impairments) were coupled to certain neurofunctional mechanisms (or impairments) in LOGG.

The biophysical mechanisms underlying BOLD signal generation are closely related to the neurochemical mechanisms measured through MRS [1]. Less efficient and weaker communication between the cerebellum and the rest of the brain was related to lower NAA and Glx as well as higher mI and mI/NAA. Lower NAA and Glx are markers of reduced neuronal health and loss of neuronal function, respectively, while elevated mI indicates neuroinflammation. Thus, a combination of poorer neuronal health, neuronal loss, and neuroinflammation likely contributed to poorer cerebellum-brain communication in LOGG. A similar interpretation extends to the association between local efficiency and NAA in the thalamus and precuneus. On a similar note, lower betweenness centrality and higher eigenvector centrality of the thalamus were related to lower NAA and creatine in LOGG (without FDR correction). Lower creatine is a marker of reduced energy metabolism. Thus, a combination of poorer neuronal health and weaker energy metabolism likely resulted in the thalamus’ pathological over-connectedness with certain brain regions (higher eigenvector centrality) and its inability to act as an effective mediator in cortical-cortical communication (lower betweenness centrality).

The local efficiency measure deserves special attention. It quantifies how well the given region communicates with other brain regions, taken together. Firstly, it was significantly lower in LOGG in the thalamus (*p* = 6.5×10^−4^), implying that the brain finds it more difficult to communicate with the thalamus in LOGG. Here it was significantly associated with disease duration as well (R = −0.94), so the brain finds it increasingly harder to communicate with the thalamus as the diseases progresses. Secondly, local efficiency was associated with NAA in all regions, having specifically a strong effect in the thalamus (R = 0.97), with the consequence of lower NAA directly implying poorer thalamo-cortical communication. Local efficiency in the cerebellum was also related to four neurochemicals, with the consequence of aberrant neurochemical levels directly implying impaired cerebellum-brain communication in LOGG. Overall, this pattern of neurochemical-neurofunctional-behavioral dysfunctional interrelationship is the central finding of this study. We believe that local efficiency in the brain must be probed further in future LOGG studies with larger samples.

Our cerebellum and thalamus results deserve attention as well. Poorer cerebellum-brain communication was related to reduced NAA and Glx and elevated mI in LOGG, while thalamus-brain communication was significantly impaired in LOGG and was also related to reduced NAA. As elaborated in the introduction section, cerebellar aberrations have been the most prominent finding in previous LOGG studies, with cerebellar atrophy [10] [11] [12] [13] [14] [15] [16], reduced NAA, and elevated mI [16]. Our study adds to this body of knowledge by demonstrating an aberrant link between cerebellum-brain communication and cerebellar NAA and mI (as well as cerebellar mI/NAA and Glx) (*p* < 0.01). On the other hand, thalamic abnormalities are lesser known in LOGG, although thalamic hypodensity [6] and lower thalamic NAA have been reported previously [9]. In fact, our study design had included the thalamus MRS VOI as an ‘at risk’ region [16]. Our current findings contribute to the growing evidence of the role of thalamic abnormalities in LOGG pathology by demonstrating an aberrant link between thalamus-brain communication and thalamic NAA (R = 0.97). Additionally, thalamic local efficiency was significantly lower in LOGG compared to controls (*p* = 6.5×10^−4^) and it was also significantly associated with disease duration (R = −0.94). Future studies must further investigate the cerebellum and thalamus in LOGG; for instance, task-based fMRI studies could be designed that target the clinical symptoms of LOGG such as ataxia, cognitive impairments and psychiatric issues, with hypotheses centered on impaired activation and task-based connectivity in the cerebellum and thalamus. Such research is needed to develop the mechanistic understanding required for building targeted treatments in the future to address these neurological, cognitive and psychiatric symptom presentations in LOGG.

Although studying group differences in graph measures was not central to this study, a secondary discussion is relevant for future research (please see Supplemental Information section S5). Finally, these fMRI measures exhibited associations with certain clinical measures. Notably, three graph measures in the thalamus were associated with disease duration, with a specifically strong association with local efficiency (R = −0.93). Longer disease duration was related to poorer communication between the thalamus and rest of the brain, implying that having the disease for longer reduces thalamo-cortical connectivity. Local efficiency was also associated with age of disease onset, with earlier age of onset resulting in poorer cerebellum-brain communication. Larger studies could use machine learning models to test whether local efficiency could be a reliable biomarker [54] and prognostic predictor of LOGG disease progression.

The primary limitation of these observations is the sample size, although our study represents a relatively large cohort (by 2 to 3 times) compared to what is normally published on this disease (N=1–3). Sample sizes in LOGG studies have remained small because of the ultra-rare nature of the disease (prevalence of about 0.0003% [23]) and corresponding challenges in patient recruitment. In fact, our study is one of the largest LOGG studies to have been conducted. We qualitatively observed that the associations seemed to be unaffected by the two LOGG disease clusters (LOTS and LOSD), implying a common mechanism coupling neurochemical and neurofunctional characteristics in them. However, we were not able to statistically ascertain this as only two LOSD patients were in our cohort. It remains to be explored if this observation and the strong associations observed in our study would hold with larger samples, and if other associations not crossing multiple comparisons correction would become stronger or weaker in larger studies. However, we believe that these observations provide a starting point for future researchers to conduct more advanced hypothesis-driven studies to further our understanding of LOGG, and ultimately to develop better diagnostic instruments and more effective treatments.

## 5. Conclusions

In this exploratory study, we sought to identify relationships between neurochemical concentrations and neurofunctional (graph) measures in LOGG. We found that reduced aggregate cerebellum-brain communication (local efficiency) was related to poorer neuronal health (NAA), neuronal loss (Glx), and neuroinflammation (mI). These findings add to the body of literature on cerebellar abnormalities in LOGG including atrophy and metabolic dysfunction. Likewise, reduced aggregate thalamus-brain communication (local efficiency) was related to poorer neuronal health (NAA), and the former was also related to longer disease duration. The thalamus is an ‘at risk’ region in LOGG based upon prior research, and our findings further attribute poor neuronal health and corresponding poor communication with the brain in this region. Future research must replicate these findings in larger cohorts, and continue to investigate neurochemical and neurofunctional abnormalities in the cerebellum and thalamus in this ultra-rare disease.

## Data Availability

Since late-onset GM2 gangliosidosis (LOGG) is an extremely rare disease, the detailed protocol approved by the IRB states that all de-identifiable imaging and clinical data (and any other study data) can be shared externally only upon IRB approval and with a data usage agreement in place.

## CRediT (Contributor Roles Taxonomy) – author contributions

**D Rangaprakash**: Conceptualization, Methodology, Software, Data Analysis, Investigation, Visualization, Writing - Original Draft, Reviewing and Editing. **Akila Weerasekera**: Data Analysis, Investigation, Writing - Reviewing and Editing. **Olivia E Rowe**: Investigation, Writing - Reviewing and Editing. **Christopher D Stephen**: Data Acquisition, Investigation, Writing - Reviewing and Editing. **Florian S Eichler**: Funding Acquisition, Data Acquisition, Investigation, Writing - Reviewing and Editing. **Robert L Barry**: Funding Acquisition, Resources, Project Administration, Conceptualization, Methodology, Data Acquisition, Investigation, Writing - Reviewing and Editing, Supervision. **Eva-Maria Ratai**: Funding Acquisition, Resources, Project Administration, Methodology, Data Acquisition, Data Analysis, Investigation, Writing - Reviewing and Editing, Supervision.

## Acknowledgments

We thank our patients and their families for participating in this research study. This study was supported by the Sanofi US Services Inc. and National Tay-Sachs & Allied Diseases Association Inc. Imaging was performed at the Athinoula A. Martinos Center for Biomedical Imaging using resources provided by the Center for Functional Neuroimaging Technologies (P41EB015896) and the Center for Mesoscale Mapping (P41EB030006), Biotechnology Resource Grants supported by the National Institute of Biomedical Imaging and Bioengineering, and the National Institutes of Health (NIH). The NIH also provided support through grants R01EB027779 and R00EB016689 (to R.L.B.). This research was also supported in part by the Athinoula A. Martinos Center for Biomedical Imaging. The content is solely the responsibility of the authors and does not necessarily represent the official views of the NIH.

## Data Availability Statement

Since LOGG is an extremely rare disease, the detailed protocol approved by the MGH IRB states that all de-identifiable imaging and clinical data (and any other study data) can be shared externally only upon IRB approval and with a data usage agreement in place.

