## Supplemental Information for "Relationship between neurochemical concentrations and neurofunctional measures in late-onset GM2 gangliosidosis"

In this document, section S1 provides MRS voxel centroids for all LOGG and control subjects, section S2 presents the relationship between MRS concentrations and node strength (graph measure), and section S3 presents the tertiary analysis on the relationship between MRS concentrations and two regional fMRI measures (fALFF and HRF). Section S4 presents the (lack of) effect of age on our results, and section S5 discusses group differences in graph measures.

### S1. MRS voxel centroids in standard space

**Table S1**. *MRS voxel centroids in the MNI space*

| | MNI Coordinates | | |
|---|---|---|---|
| **Subject** | **Cerebellum** | **Thalamus** | **Parietal** |
| LOGG-01 | -2, -64, -28 | 14, -16, 4 | 2, -48, 48 |
| LOGG-02 | 0, -62, -28 | 10, -16, 4 | 0, -52, 44 |
| LOGG-03 | 0, -58, -24 | 14, -18, 4 | 2, -42, 44 |
| LOGG-04 | 0, -56, -26 | 16, -16, 4 | 0, -42, 50 |
| LOGG-05 | 0, -68, -30 | 18, -16, 4 | 0, -44, 48 |
| LOGG-06 | 0, -64, -28 | 16, -16, 8 | 0, -40, 46 |
| LOGG-07 | 0, -62, -28 | 18, -16, 8 | 2, -42, 48 |
| control-01 | 0, -62, -30 | 16, -16, 6 | 2, -46, 50 |
| control-02 | 0, -62, -30 | 14, -16, 6 | -2, -40, 48 |
| control-03 | 0, -60, -28 | 10, -18, 6 | 2, -46, 56 |
| control-04 | 0, -64, -28 | 14, -18, 6 | 2, -42, 48 |
| control-05 | 0, -62, -28 | 14, -16, 8 | 2, -44, 54 |
| control-06 | 0, -64, -28 | 18, -14, 6 | 2, -46, 52 |
| control-07 | 0, -64, -32 | 18, -16, 4 | 4, -48, 48 |

### S2. Relationship between MRS concentrations and node strength

We observed no significant associations between neurochemicals and node strength in any region (*p*<0.05, FDR corrected), although node strength was associated with NAA, mI, mI/NAA and Glx in the cerebellum without multiple comparisons correction (**Table S2**). These observations go hand-in-hand with local efficiency findings in the cerebellum (**Table 2**, main text) and direct us towards the inference that impaired aggregate cerebellum-brain communication is related to reduced NAA and Glx and elevated mI and mI/NAA in LOGG.

**Table S2.** *Significant associations between MRS concentrations and node strength (graph measure), without multiple comparisons correction. Node strength was associated with NAA, mI, Glx and mI/NAA. NAA = N-acetyl aspartate, mI = myo-inositol, Glx = glutamate + glutamine.*

| | Graph measure | MRS measure | R-value | $R^2$ value | p-value |
|---|---|---|---|---|---|
| | **Cerebellum** | | | | |
| 1 | Node strength | NAA | 0.77 | 0.58 | 0.0428 [a] |
| 2 | | mI | −0.86 | 0.74 | 0.0133 [a] |
| 3 | | mI / NAA | −0.79 | 0.62 | 0.0353 [a] |
| 4 | | Glx | 0.82 | 0.68 | 0.0226 [a] |

[a] *not significant after multiple comparisons correction*

### S3. Relationship between MRS concentrations and two other regional fMRI measures

In this study, our primary focus was on nodal graph measures that quantify the aggregate communication between the MRS ROIs and rest of the brain. However, given the paucity of imaging studies in LOGG, we additionally performed certain tertiary analyses with the hope that these supplementary observations would guide further hypothesis driven LOGG fMRI studies in the future. These tertiary observations are not part of the main conclusions of this study.

As noted in the main text, various time series analysis techniques could be employed to understand rich fMRI time series data to gain mechanistic insights about neurological disorders such as LOGG. Among such approaches, we present observations from two regional fMRI measures viz. fractional amplitude of low frequency fluctuations (fALFF) and hemodynamic response function (HRF). Both these techniques provide one value for each MRS ROI, enabling us to directly correlate them with the MRS concentrations.

#### S3.1. Associations between MRS concentrations and the strength of low frequency BOLD fluctuations (fALFF)

FALFF [1] is a measure of the contribution of low frequency (0.01–0.10 Hz) BOLD fluctuations relative to the entire frequency range. FALFF can be considered as the resting-state analogue of 'activation', that is, a measure of BOLD signal strength. FALFF was computed in the frequency domain (upon applying Fourier transform on the time series) as the ratio of the fMRI signal power in the low frequency band (0.01–0.1 Hz) to the total signal power in the entire frequency range (which is 0–0.25 Hz for our data with TR=2s) [2]. Since most of the neuronal signal correlates reside in this low frequency band in fMRI, and signal outside this band is typically considered as dominated by noise, this ratio represented the strength of BOLD signal in resting state (analogous to 'activation' in task fMRI). One fALFF value was obtained per time series. Since fALFF required unfiltered data, we first obtained mean MRS ROI time series from unfiltered deconvolved fMRI data and then computed fALFF for each of the 3 ROIs in all subjects. The fALFF values in each ROI were then correlated with each of the six MRS concentrations (Cho, Cr, mI, Glx, NAA and mI/NAA) in the corresponding MRS ROI within the LOGG group, and the statistics were controlled for multiple comparisons using the false discovery rate (FDR) method ($p<0.05$). Separately, group differences in fALFF values were also assessed.

FALFF was lower in LOGG compared to controls ($p = 0.037$, $T = 2.35$) only in the cerebellum, with a decent effect size (Cohen's $d = 1.25$). FALFF was not associated with any of the six neurochemicals in any of the MRS ROIs.

Finally, we assessed the association between fALFF values (in the three MRS voxels) and clinical variables ($p<0.05$, FDR corrected for multiple comparisons). Specifically, we considered the following clinical measures as mentioned in the main text: age at disease onset, disease duration, LOGG severity, Epworth sleepiness scale, Pittsburgh sleep quality index (PSQI), eating assessment tool (EAT10), brief ataxia rating scale (BARS), cerebellar cognitive affective syndrome scale (CCAS), Friedreich ataxia rating scale (FARS), and semiquantitative assessment of cerebellar ataxia (SARA).

FALFF in the cerebellum was significantly positively associated with disease duration (R = 0.96, $R^2 = 0.92$, $p = 6.1\times10^{-4}$) (**Table S3**). Given the central role of cerebellum in LOGG pathology [3], it is interesting that fALFF and disease duration were highly related, with longer disease duration related to higher fALFF values. The regression fit (**Figure S1**) was fully robust: there were no influential observations; the min and max p-values with leave-one-out regression were 0.0002 and 0.0077 respectively; the residuals were homoscedastic ($p = 0.69$).

**Table S3.** *Significant associations between fALFF and clinical variables.*

| | fMRI measure | Behavioral measure | R-value | $R^2$ value | p-value |
|---|---|---|---|---|---|
| | **Cerebellum** | | | | |
| 1 | fALFF | Disease duration | 0.96 | 0.92 | $6.1 \times 10^{-4}$ |

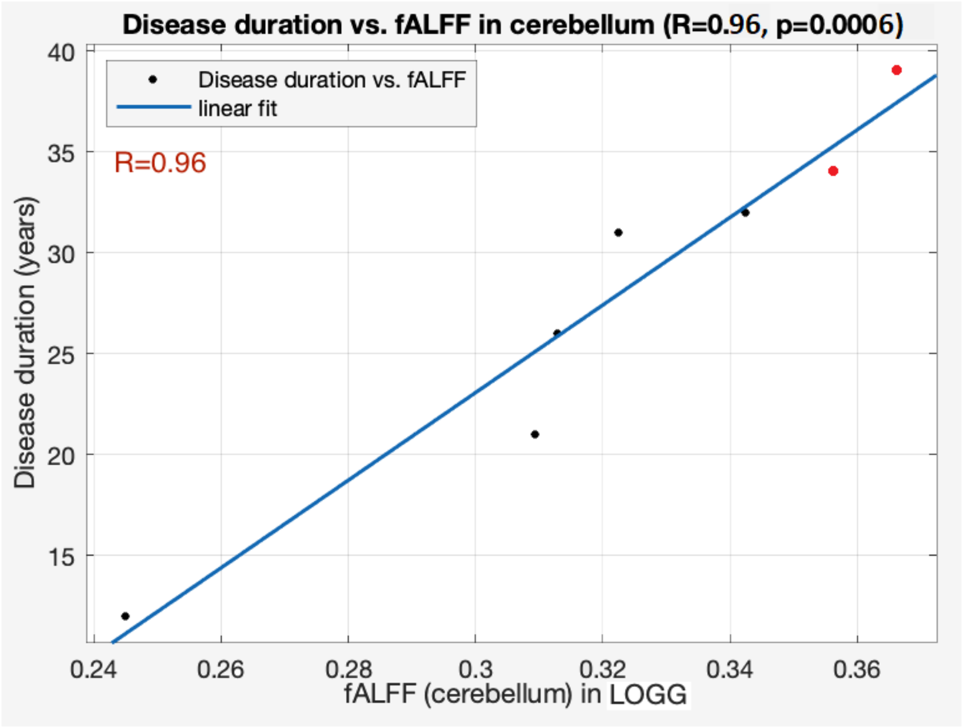


**Figure S1**. *Significant association between disease duration and fALFF in the cerebellum (R = 0.96, $R^2$ = 0.92, p = 0.00061). The two LOSD patients are visible as red points; it is evident that this association was not largely driven by differences between LOTS and LOSD.*

#### S3.2. Associations between MRS concentrations and neurovascular coupling (HRF)

Another fMRI-derived measure considered was the hemodynamic response function (HRF), which represents neurovascular coupling. It is the bridge between the latent neural activity and the measured BOLD fMRI signal, and is tightly related to local neurochemistry [4] [5], motivating us to perform a supplemental MRS-HRF analysis here. Specifically, we performed an exploratory analysis assessing the relationship between the six neurochemicals (Cho, Cr, mI, Glx, NAA and mI/NAA) and HRF parameters in the same MRS ROI.

While fALFF measures BOLD signal strength, it does not untangle the latent neuronal signal and neurovascular coupling that make up the BOLD signal. The HRF represents neurovascular coupling, and here we studied the relationship between MRS concentrations and the HRF within the LOGG group to better understand LOGG neurobiology. The deconvolution step (described under pre-processing in the main text) provided us HRF estimates within each voxel. HRF shape is typically characterized with three parameters: response height (RH), full-width-at-half-maximum (FWHM) and time-to-peak (TTP) [6] [7]. RH is the amplitude of neurovascular coupling, FWHM is related to BOLD response duration, and TTP is the BOLD response latency. With FWHM having a range of 1.1–1.9s and TTP having a range of 2.5–6.5s [6], our data (TR=2s) was considered to not have sufficient temporal resolution for FWHM/TTP analysis; thus, only RH was examined. HRF RH values within the gray matter of each MRS voxel were averaged to provide a single RH value for each ROI in each subject. Associations between RH values and MRS concentrations within the LOGG group were obtained ($p<0.05$, FDR corrected). Separately, group differences in RH values were also assessed.

HRF RH was not significantly different between groups in any of the three MRS voxels, indicating that neurovascular coupling is likely undisturbed in LOGG in these regions. Without multiple comparisons correction, RH in the thalamus was associated with disease duration ($R = -0.77$, $R^2 = 0.59$, $p = 0.045$), with lower RH related to longer disease duration.

RH was significantly negatively associated with creatine in the thalamus ($R = -0.96$, $R^2 = 0.92$, $p = 5.5\times10^{-4}$) (**Table S4**). Without multiple comparisons correction, RH was also associated with NAA in the thalamus and choline in the precuneus (see Table 4). Across all regions, higher amplitude of neurovascular coupling was related to reduced neurochemical concentrations. The RH vs. creatine regression fit (**Figure S2**) was fully robust: there were no influential observations;

the min and max p-values with leave-one-out regression were 0.0004 and 0.016 respectively; the residuals were homoscedastic ($p$ = 0.69).

**Table S4.** *Significant associations between MRS concentrations and HRF response height. The association between HRF response height and creatine in the thalamus and with choline in the precuneus (in black) survived multiple comparisons correction (FDR method), while the other association (in gray) did not. Choline = GPC+PCh (total glycerophosphol-choline + phospho-choline); creatine = Cr+PCr (total creatine + phosphocreatine); NAA = N-acetyl aspartate.*

| | fMRI measure | MRS measure | R-value | $R^2$ value | p-value |
|---|---|---|---|---|---|
| | **Thalamus** | | | | |
| 1 | HRF response height | Creatine | −0.96 | 0.92 | $5.7 \times 10^{-4}$ |
| 2 | HRF response height | NAA | −0.82 | 0.67 | 0.0246 [a] |
| | **Precuneus** | | | | |
| 3 | HRF response height | Choline | −0.85 | 0.73 | 0.0150 [a] |

[a] *not significant after multiple comparisons correction*

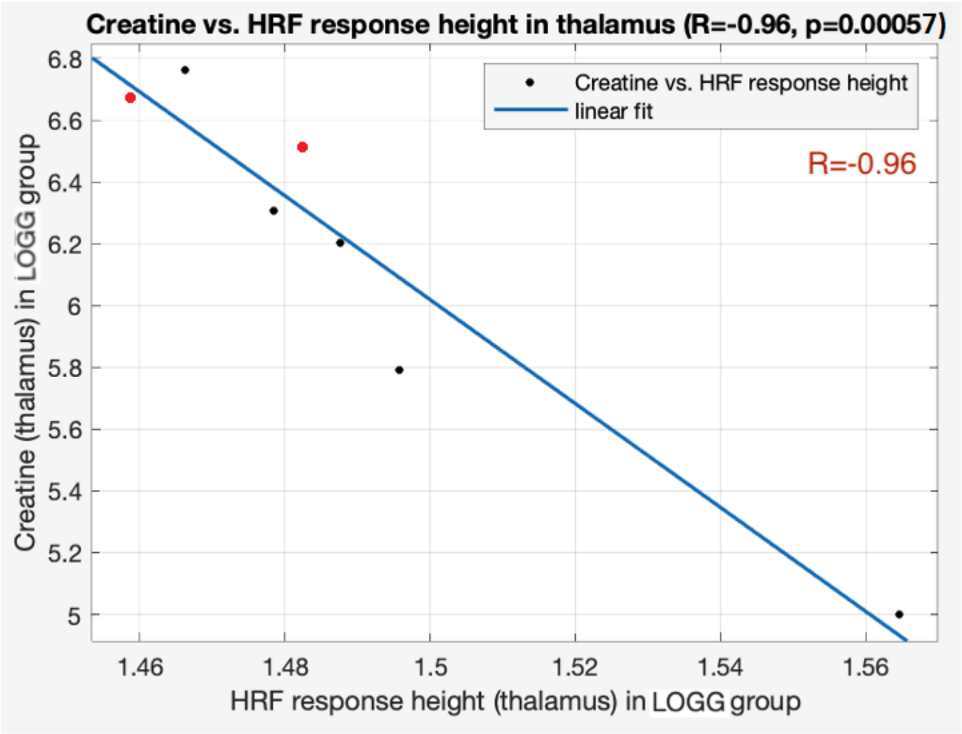

**Figure S2**. *Significant association between creatine and. HRF response height in the thalamus ($R = -0.96$, $R^2 = 0.92$, $p = 0.00057$). The two LOSD patients are visible as red points.*

In summary, fALFF was not significantly associated with MRS concentrations but was significantly lower in LOGG and highly associated with disease duration (both in the cerebellum ROI), while HRF RH was significantly associated with choline in the precuneus (and with creatine in thalamus without correction), but not different between groups or associated with clinical variables. Higher neurovascular coupling strength was related to lower neurochemical concentrations, implying that reduced creatine or choline was perhaps resulting in elevated neurovascular response in these regions in LOGG.

Longer disease duration was related to overall higher thalamic BOLD signal strength and poorer communication between the thalamus and the rest of the brain (graph analysis results presented in main text), implying that having the disease for longer causes more thalamic activity but reduces thalamo-cortical connectivity. Higher BOLD signal strength can coexist with poorer connectivity; for instance, multiple studies have reported higher activations in task fMRI studies coexisting with lower connectivity between activated regions indicating asynchronous pathological neuronal overdrive [8] in these disorders.

### S4. The effect of age on MRS-fMRI relationship in LOGG

Age was not significantly different between LOGG and control groups ($p = 0.95$). The LOGG group's age range was 22 to 62 years. The LOGG group was comprised of five participants from the LOTS variant and two from the LOSD variant. The LOSD participants were older (median age = 57 years) than the LOTS participants (median age = 36 years). With such a small sample of LOSD patients, we were unable to perform group comparisons (as mentioned in the main text). However, we tested the effect of age on our results by (i) testing for significant correlations of age with MRS concentrations and fMRI measures, and (ii) repeating our analyses (only significant results) with age as a covariate of no interest. The findings are presented next.

After multiple comparisons correction ($p<0.05$, FDR method), age was not significantly correlated with any of the MRS concentrations or graph measures. Without correction ($p<0.05$), in the cerebellum, age correlated with NAA (R = 0.78, $p$ = 0.037), mI (R = –0.79, $p$ = 0.036) and Glx (R = 0.76, $p$ = 0.047), as well as with local efficiency at a trend level (R = –0.69, $p$ = 0.083).

In the main text, we reported an association between these measures (local efficiency and neurochemicals) in the cerebellum (Table 2). To account for the effect of age, we repeated the analysis with age as a covariate. The results did not differ, with these associations remaining significant after accounting for age: local efficiency vs. NAA (R = 0.83, $p$ = 0.022), local efficiency vs. mI (R = 0.93, $p$ = 0.003), local efficiency vs. Glx (R = 0.84, $p$ = 0.018). This shows that, despite age correlating with these measures in the cerebellum (without multiple comparisons correction), age had a proportional effect on both neurochemicals and local efficiency such that the association between the measures remained significant.

We also repeated the analysis with other significant results from the main text (wherein the individual measures did not correlate with age). All the associations presented in the main text remained significant even after using the age covariate. Specifically, all associations presented in Table 2 were significant ($p<0.05$, FDR corrected, same as in main text), all associations in Table 3 were significant ($p<0.05$, uncorrected, same as in main text), and all associations in Table 4 were significant ($p<0.05$, FDR corrected, same as in main text) except eigenvector centrality vs. disease duration that was significant without FDR correction (R = 0.82, $p$ = 0.045). In other words, all the results presented in the main text remained significant, except for the eigenvector centrality vs. disease duration association that was significant with an uncorrected threshold but not with FDR correction. This invariance of our associations with age is also reflected in most cases in Figures 1–3 of the main text, with the LOSD patients in red visually not distinct from the LOTS patients' cluster.

### S5. A discussion on group differences in graph measures

In the results section of the main text (section 3.1), we presented group differences in graph measures as follows:

With group comparisons (LOGG vs. controls), local efficiency was significantly lower in LOGG compared to controls in the thalamus ($p = 6.5\times10^{-4}$, $T$ = 4.57, Cohen's $d$ = 2.44), indicative of poorer thalamus-brain communication in LOGG. Without multiple comparisons correction, local efficiency was also significantly lower in LOGG in the precuneus ($p$ = 0.034, $T$ = 2.39, Cohen's $d$

= 1.28). Local efficiency was not different in the cerebellum ($p$ = 0.35). Other graph measures did not differ between the groups in the three MRS ROIs ($p$>0.05).

Although studying group differences in graph measures was not central to this study, a secondary discussion is provided here in the interest of future research. Firstly, thalamus-brain communication (local efficiency) was significantly impaired in LOGG ($p = 6.5\times10^{-4}$), but local efficiency does not inform us specifically which subset of connections were most affected. Comparing the 258 FC values between thalamus and rest of the brain (LOGG vs. controls), no significant differences were found with FDR correction, but 7 connections were seen without multiple comparisons correction ($p$<0.05) in default mode (3 connections), visual, ventral attention, somatomotor, and cingulo-opercular task control networks (1 connection each). (Please refer to [9] to learn more about these networks.) Although these observations are not robust given our sample size, we believe our study has provided enough motivation to probe thalamo-cortical connectivity impairments thoroughly in the future. Next, neither local efficiency nor any other graph measure was impaired in LOGG in the cerebellum, implying that aggregate cerebellum-brain communication was not impaired enough in LOGG. However, this does not imply that certain specific subset of individual connections between cerebellum and rest of the brain were not impaired, which future studies need to explore. This also does not discount the relatively large body of literature on anatomical aberrations in the cerebellum of LOGG patients (such as cerebellar atrophy). Although cerebellum-brain FC was not significantly different with FDR connection, five connections were each seen without correction ($p$<0.05) in somatomotor, default mode, visual, ventral attention and frontoparietal task control networks. Likewise, four precuneus-brain connections were seen without correction ($p$<0.05) in somatomotor, default mode, dorsal attention, and frontoparietal task control networks. These tertiary observations, although not central to our study's aim of assessing MRS-fMRI relationship in LOGG, are sufficiently relevant to mention as motivation for future studies.
